# Investigating potential myopia risk factors, including chronotype, in Estonian adolescents

**DOI:** 10.1101/2023.09.12.23294974

**Authors:** Teele Palumaa, Delis Linntam, Reili Rebane, Marika Tammaru, Kadi Palumaa

## Abstract

**Purpose:** To evaluate chronotype, lifestyle factors, and parental myopia in relation to myopia, and characterise the effect of cycloplegia on spherical equivalent refraction (SER) in Estonian secondary school students.

**Methods:** One hundred twenty-three students aged between 15 and 17 years from three secondary schools in Estonia participated in the study. They underwent a comprehensive ocular examination, including cycloplegic refraction and ocular biometry. Chronotype was evaluated with the Morningness–Eveningness Questionnaire. Participants also completed a questionnaire about their daily activities, including time spent outdoors, time spent on near-work and mid-working distance activities, and parental myopia. Myopia was defined as cycloplegic SER ≤–0.50 D. Logistic regression analysis was performed to assess the association of the studied factors with myopia.

**Results:** In a multivariate regression model, having two myopic parents was associated with higher odds of myopia (OR 3.78, 95% CI 1.15–12.42). We found no association between myopia and chronotype. Notably, time spent outdoors, and doing near-work and mid-working distance activities, did not affect the likelihood of having myopia. Non-cycloplegic SER was significantly more myopic than cycloplegic SER on average by 0.86 D (*p* < 0.0001, Wilcoxon matched-pairs signed rank test).

**Conclusion:** Consistent with previous reports, we identified parental myopia as a myopia risk factor. Chronotype was not associated with myopia in our study sample. Interestingly, there was no association between myopia and time spent outdoors or near work. Using non-cycloplegic refraction would lead to a significant overestimation of myopia.

## Introduction

The prevalence of myopia, or short-sightedness, has increased markedly over the last decades [1]. Should the current trend continue, approximately half of the global population will be myopic by 2050 [1]. Myopia risk factors have been studied extensively worldwide. It has emerged that the risk of developing myopia is reduced in children who spend more time outdoors [2–5] and increased with more near-work [6], and computer use [7].

Several recent studies have raised the possibility that myopia is associated with circadian rhythms. Genes associated with refractive errors in genome-wide studies have an increased representation of genes controlling circadian rhythms [8]. It is also known that axial length and choroidal thickness show diurnal rhythmicity in humans [9, 10] and chicks [11, 12]. Furthermore, inducing myopia in chicks with form deprivation abolishes this circadian rhythmicity in axial length [11].

There is also evidence to suggest that short-sightedness may be linked to sleep. For example, myopic children have been reported to sleep later than children with no myopia [13– 16], and to have worse sleep quality [17, 13]. However, the data in the literature is inconsistent and several studies have found no associations between myopia and sleep quality [18] and bedtime [19, 20]. Interestingly, though, poor sleep quality is associated with evening chronotype, or one’s late sleep-wake timing preference [21–23], raising the possibility that late chronotype may be the link between myopia and poor sleep quality. Chronotype has been studied in the context of myopia in a couple of studies, and no definitive associations have been found. In one report, the circadian phases of myopes and non-myopes did not differ [24] and in another, self-report of a “morning” or “evening” type was not associated with myopia progression [15].

Some discrepancies between the findings of the studies listed above might be related to whether myopia was self-reported [16], defined with cycloplegic [14, 19, 18, 20, 15] or non-cycloplegic [13, 24] refraction measurements. It is known that refraction measurements are more myopic without cycloplegia [25, 26], which may lead to overestimating the number of myopes in studies using non-cycloplegic refraction to determine myopia.

In this study, we assessed the effect of chronotype, parental myopia status, and lifestyle factors on myopia in Estonian adolescents. We also analysed the difference between cycloplegic and non-cycloplegic spherical equivalent refraction (SER) measurements to estimate its influence on assessing myopia prevalence in our study population.

## Methods

The study was conducted at the East Tallinn Central Hospital Eye Clinic in 2019. It adhered to the principles of the Declaration of Helsinki and was approved by the Tallinn Medical Research Ethics Committee. Form 10 students from three public schools in Tallinn were invited to participate, and a total of 123 students were recruited. Written informed consent was obtained from the participants and their parents or legal guardians. All participants were European Caucasians. The schools differed slightly in their education profile, putting more emphasis on science, arts, or sports.

Participants underwent a comprehensive ocular examination, including non-cycloplegic and cycloplegic refraction, ocular biometry and biomicroscopy. Cycloplegia was achieved by applying two rounds of 1% cyclopentolate and 0.5% tropicamide with a 5-minute interval, as per [3]. Cycloplegia was evaluated 40 minutes after the application of the first round and considered adequate when the pupil was ≥6 mm in diameter and not responsive to light. Ocular refraction was measured with HandyRef-K autorefractor (Nidek, Tokyo, Japan), axial length and anterior corneal curvature with IOLMaster 500 (Zeiss, Oberkochen, Germany). SER was calculated as sphere + 1/2 cylinder, and participants’ SER was defined as the mean SER of the left and right eyes. Myopia was defined as cycloplegic SER≤–0.50 dioptres (D), high myopia as SER ≤–6.00 D, astigmatism as ≤–1.00 dioptre cylinder and hyperopia as SER ≥ 2.00 D.

Participants filled out a questionnaire about daily activities, including time spent outdoors, doing near work and mid-working distance activities, separately for weekdays and free days. Total time spent on each type of activity was calculated per week. Near-work activities were defined as reading, writing, and using a smartphone. Mid-working distance activities were defined as watching TV, working with a computer, and playing video games. The weekly total number of hours spent outdoors, doing near work and mid-working distance activities were calculated and used in the analyses. Chronotype was evaluated with the Morningness-Eveningness Questionnaire (MEQ) [27]. MEQ was translated from English into Estonian and pilot-tested on a separate cohort of thirty 16-17-year-olds to eliminate any misunderstandings in the wording. The questionnaire also contained questions about parental myopia.

Statistical analyses were performed using Stata Software for Statistics and Data Science (version 14.2). Graphs were produced in Prism (version 9, GraphPad). Continuous variables were expressed as mean or median when appropriate, and qualitative variables were summarised as frequency and percentage. Wilcoxon signed-rank test was used to compare the SER before and after cycloplegia and Spearman rho was calculated to characterise correlations between ocular parameters. Variables associated with myopia were assessed with multivariate logistic regression analysis, and results presented as odds ratios (OR) with their 95% confidence interval (CI). A two-tailed *p* value of <0.05 was considered statistically significant.

## Results

A total of 123 students participated in the study (57% female), and the majority were 16 years old (71%). 18% reported having two myopic parents, and 41% had one myopic parent. Half of the participants were from the science-oriented school, 28% from the arts-oriented and 22% from the sports-oriented school (see Table 1 for a demographic characterisation). The prevalence of myopia in our study population was 30.9%, high myopia was present in 2.4%, hyperopia in 3.3% and astigmatism in 11.4%.

**Table 1.** Demographic characteristics of the study population. The demographics are characterised for the entire study population. The number and proportion of myopes in each demographic group are presented.

|  | All participants |  | Myopes |  |
| --- | --- | --- | --- | --- |
|  | N | % | N | % |
| **Total** | 123 | 100 | 38 | 30.9 |
| ***Characteristic*** |  |  |  |  |
| **Sex** |  |  |  |  |
| Female | 70 | 56.9 | 24 | 34.3 |
| Male | 53 | 43.1 | 14 | 26.4 |
| **Age** |  |  |  |  |
| 15 years | 5 | 4.1 | 1 | 20.0 |
| 16 years | 87 | 70.7 | 26 | 29.9 |
| 17 years | 31 | 25.2 | 11 | 35.5 |
| **Parental myopia** |  |  |  |  |
| None | 43 | 35.0 | 8 | 18.6 |
| One myopic parent | 58 | 47.2 | 19 | 32.8 |
| Both parents myopic | 22 | 17.9 | 11 | 50.0 |
| **School** |  |  |  |  |
| Science | 61 | 49.6 | 27 | 42.3 |
| Arts | 35 | 28.5 | 8 | 22.9 |
| Sports | 27 | 22.0 | 3 | 11.1 |

The average time spent outdoors per week was 16.5 hours for both non-myopes and myopes (Fig. 1A). Myopes spent slightly more time on near-work activities, 54.4 hours, compared to 52.0 hours per week in non-myopes (Fig. 1B). The average time spent on mid-working distance activities was 21.3 hours for non-myopes and 17.7 hours for myopes (Fig. 1C). In MEQ, a larger score indicates an earlier chronotype [27]. In our study sample, the MEQ score was 46.7 for non-myopes and 48.8 for myopes (Fig. 1D).

**Figure 1.**
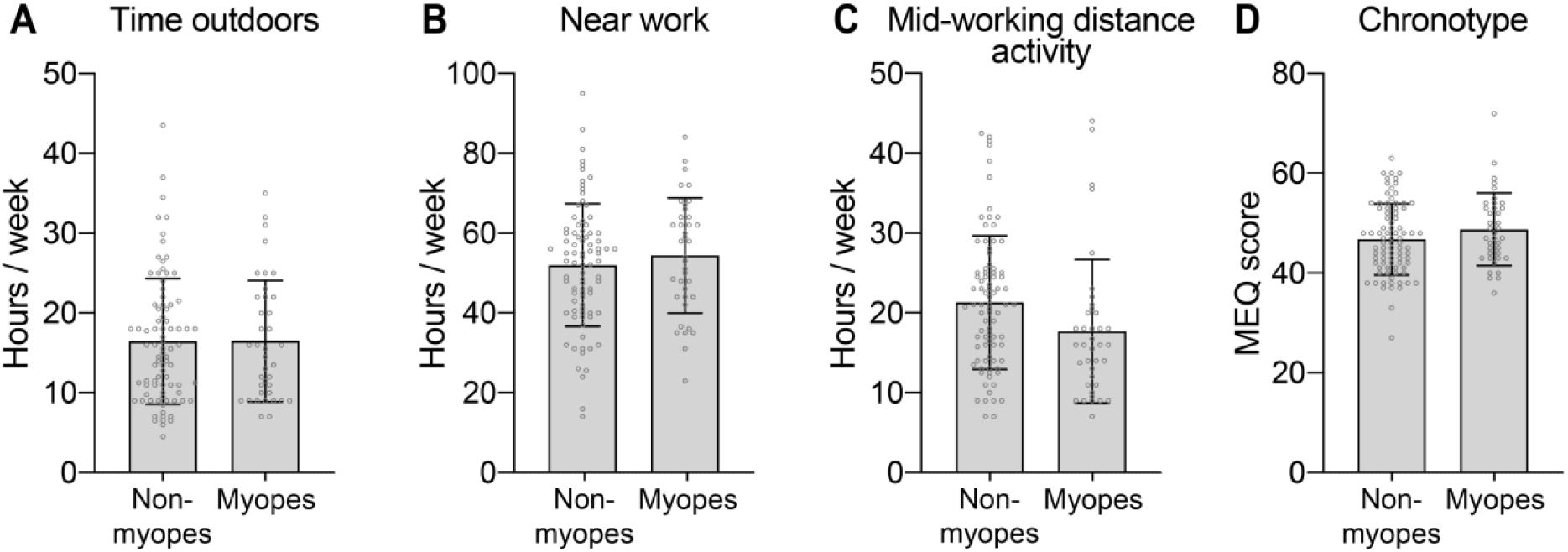
Distribution of factors analysed in non-myopes and myopes. (A) Time spent outdoors, (B) on near work and (C) mid-working distance activities in non-myopes and myopes. (D) Distribution of chronotype, measured with MEQ, in non-myopes and myopes. Data are presented as mean ± SD. MEQ, Morningness-Eveningness Questionnaire; SD, standard deviation.

Multivariate logistic regression analyses of the factors studied (Table 2) revealed that participants with two myopic parents were 4.3 times more likely to have myopia (95% CI 1.19 – 15.74). Subjects attending the sports-oriented school had lower odds of myopia than participants attending the science-oriented school (OR 0.12, 95% CI 0.03 – 0.51). We found no significant association between chronotype and refractive status. Time spent outdoors, on near work and mid-working distance activities were not associated with myopia in our study.

**Table 2.** Effect of demographic and lifestyle factors and chronotype on myopia status. A multivariate logistic regression model was fitted with myopia status as the dependent variable. Sex, parental myopia, school, chronotype tercile, outdoor time tercile, near work tercile and mid-working distance activity time tercile were included as independent variables. A *p* value of <0.05 was considered statistically significant and marked in bold. CI, confidence interval; MEQ, Morningness-Eveningness Questionnaire.

| Characteristic | Odds ratio | p value | 95% CI |
| --- | --- | --- | --- |
| **Sex** |  |  |  |
| Female | 1.00 |  |  |
| Male | 1.83 | 0.213 | 0.71 – 4.76 |
| **Parental myopia** |  |  |  |
| None | 1.00 |  |  |
| One myopic parent | 1.50 | 0.459 | 0.52 – 4.33 |
| Both parents myopic | 4.32 | **0.026** | 1.19 – 15.74 |
| **School** |  |  |  |
| Science | 1.00 |  |  |
| Arts | 0.41 | 0.103 | 0.14 – 1.20 |
| Sports | 0.12 | **0.004** | 0.03 – 0.51 |
| **Chronotype (MEQ score)** |  |  |  |
| I tercile (evening types) | 1.00 |  |  |
| II tercile | 2.29 | 0.158 | 0.73 – 7.26 |
| III tercile (morning types) | 2.69 | 0.074 | 0.91 – 7.98 |
| **Time spent outdoors (hours per week)** |  |  |  |
| I tercile (4.5-11.0) | 1.00 |  |  |
| II tercile (11.25-17.75) | 1.17 | 0.794 | 0.35 – 3.90 |
| III tercile (18.0-43.5) | 1.00 | 0.996 | 0.32 – 3.14 |
| **Time spent on near work (hours per week)** |  |  |  |
| I tercile (14.0-45.0) | 1.00 |  |  |
| II tercile (46.0-59.0) | 0.58 | 0.363 | 0.18 – 1.86 |
| III tercile (60.0-95.0) | 1.00 | 0.996 | 0.32 – 3.15 |
| **Time spent on middle-distance work (hours per week)** |  |  |  |
| I tercile (3.0-15.0) | 1.00 |  |  |
| II tercile (15.5-24.5) | 1.88 | 0.270 | 0.61 – 5.76 |
| III tercile (25.0-68.0) | 0.97 | 0.961 | 0.30 – 3.19 |

The SER values displayed a left-skewed distribution in our study population with the median cycloplegic SER +0.25 D and mean SER –0.38 D (Fig. 2A). Myopia exhibited both an axial and corneal component as more myopic refraction was correlated with both longer axial length (*r* = –0.76, Fig. 2B), and smaller radius of corneal curvature, or steeper corneas (*r* = 0.17, Fig. 2C).

**Figure 2.**
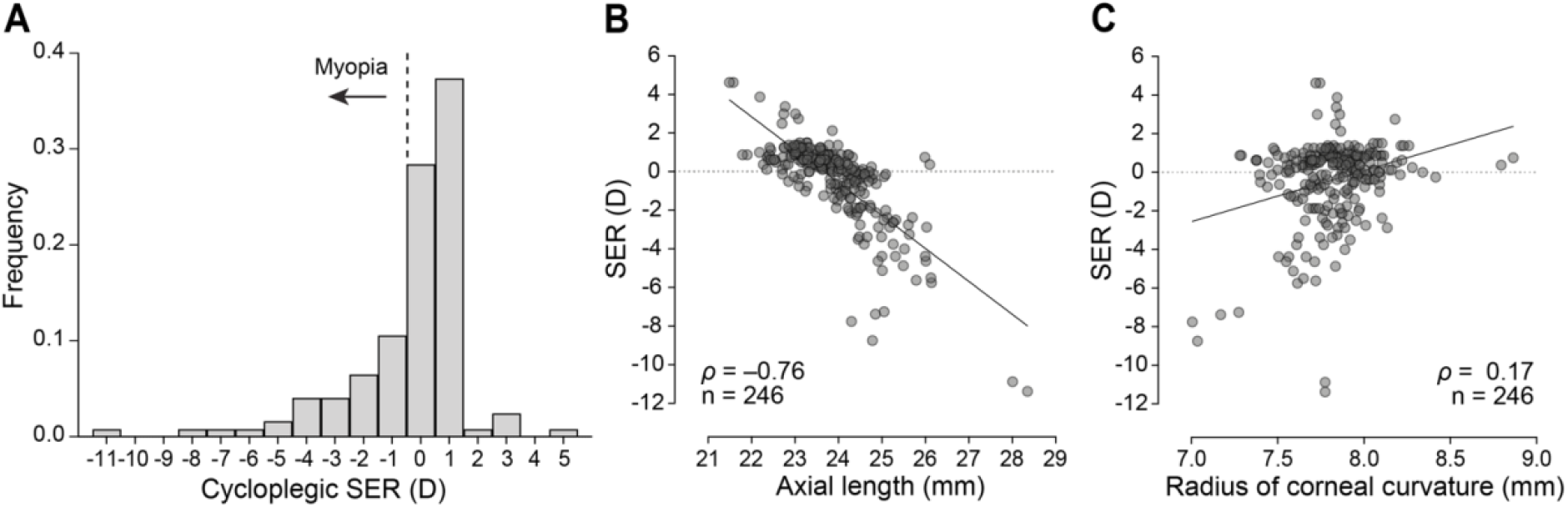
Characterisation of SER of the study population. (A) Distribution of the participants’ mean cycloplegic SER grouped into one dioptre bins. Correlations between (B) SER and axial length, and (C) SER and radius of corneal curvature in individual eyes (Spearman correlation, individual values fitted with simple linear regression to illustrate the relationship). SER, spherical equivalent refraction; D, dioptres.

The SER with and without cycloplegia were significantly different, non-cycloplegic SER was, on average, 0.89 D more myopic than cycloplegic SER (mean non-cycloplegic SER – 1.27 D, mean cycloplegic SER –0.38 D; median non-cycloplegic SER –0.88 D, median cycloplegic SER +0.25 D, *p* < 0.0001, Wilcoxon matched-pairs signed rank test, Fig. 3A). Non-cycloplegic SER was ≤–0.50 D in 83 participants (67% of the study population) while cycloplegic SER was ≤–0.50 D in 38 participants (31% of the study population). Therefore, had we defined myopia based on non-cycloplegic SER, we would have overestimated the prevalence of myopia by more than two times. The non-cycloplegic and cycloplegic SER were highly correlated (*r* = 0.74, Spearman correlation, Fig. 3B).

**Figure 3.**
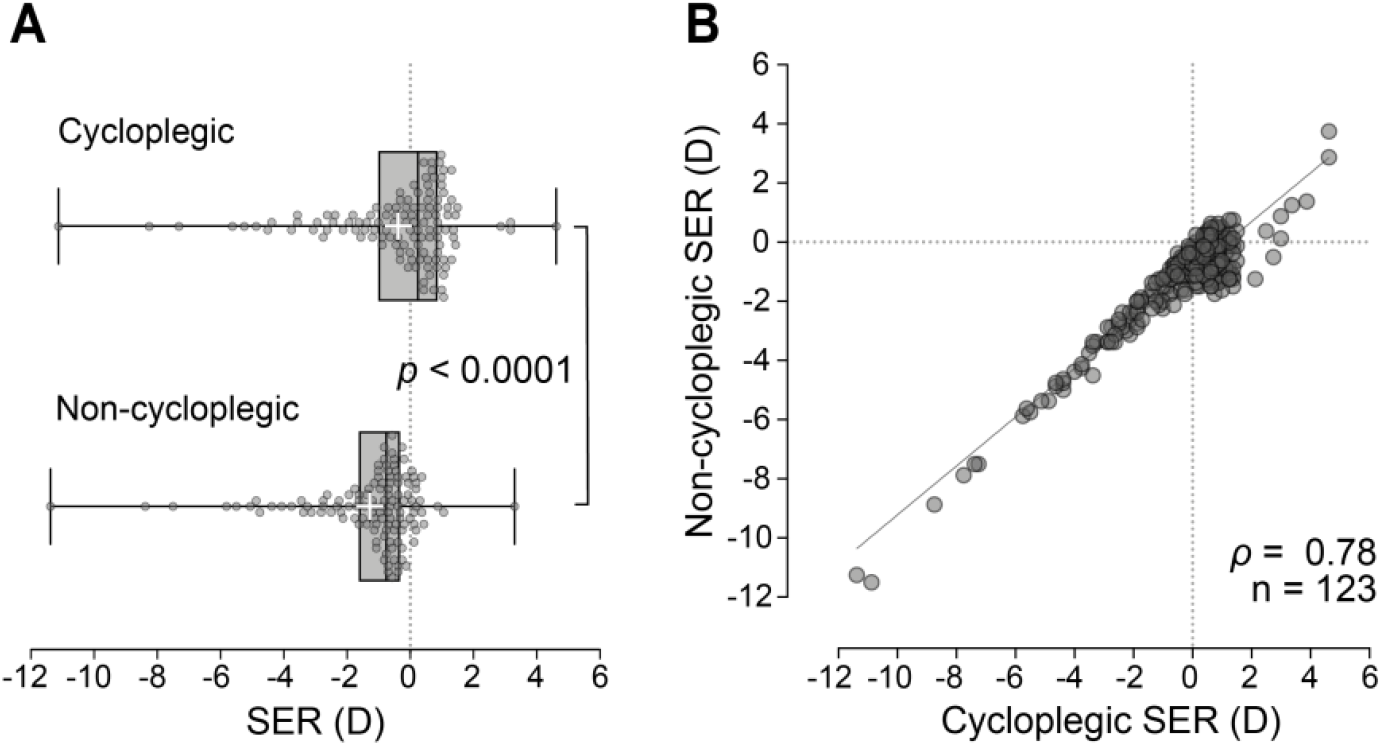
Effect of cycloplegia on SER. (A) Difference between cycloplegic and non-cycloplegic SER (*p* < 0.0001, Wilcoxon matched-pairs signed rank test; boxplot presents median with minimum and maximum values, “+” denotes the mean, individual participant values presented). (B) Correlation between cycloplegic and non-cycloplegic SER (Spearman correlation, individual values fitted with simple linear regression to illustrate the relationship). SER, spherical equivalent refraction; D, dioptres.

## Discussion

This is the first study to investigate myopia risk factors in Estonia. Chronotype, assessed with MEQ, was not associated with myopia in our sample. While later bedtime has been associated with myopia [13–16], a link between chronotype and myopia has not been established. In their study, Flanagan et al. [24] found that myopes had higher melatonin levels at all time points studied, while the circadian parameters, i.e. dim-light melatonin onset, phase of circadian activity determined with actigraphy, and MEQ score did not differ between myopes and non-myopes. It should be mentioned that non-cycloplegic refraction was used to determine myopia [24]. Furthermore, self-reported “morning” or “evening” type was also not associated with myopia progression in a study by Lee et al. [15]. Larger studies are needed to further evaluate the potential link between human circadian rhythms and refractive errors.

In agreement with other investigations [28], we determined parental myopia as a myopia risk factor. While increased time spent outdoors has been convincingly shown to protect from myopia development in multiple studies [2–5], we did not see such a correlation in our study population. The reason behind this observation may be our limited sample size and the use of a questionnaire to measure time spent outdoors. It has been demonstrated that a daily activities diary correlates better with time spent outdoors evaluated by wearable light meters than a questionnaire [29]. However, this is not the first study to report no association between time outdoors and myopia in adolescents. Schmid et al. ^37^[30] found no differences in the time spent in sunlight between 17 to 25-year-old myopes and non-myopes. Lu et al. [31] reported no differences in self-reported time spent outdoors and myopia status in Singaporean adolescents (mean age 14.7 years). Furthermore, Hagen et al. [32] saw no differences in reported outdoor time between myopes and non-myopes in 16 to 19-year-old Norwegian adolescents. Our study participants were aged between 15 to 17 years, and it may be that outdoor time at a younger age, which is more critical for refractive development, confers a protective effect, and outdoor time during adolescence may not be reflective of the previous outdoor exposure.

We assessed the SER before and after cycloplegia and determined an average difference of –0.89 D, with the non-cycloplegic values being more myopic. This is similar to a previous study reporting a –0.63 D difference [26]. Had we used non-cycloplegic SER values, we would have overestimated the prevalence of myopia in our study population by more than two-fold. This highlights the striking difference in the measured non-cycloplegic and cycloplegic autorefraction values and emphasises the need to use cycloplegia for determining refractive status, which may be one underlying cause of conflicting results in the literature.

In conclusion, this is the first study evaluating myopia risk factors in Estonia. In agreement with published work, we found that parental myopia was a risk factor for myopia. Decreased time spent outdoors and increased time on near-work activities were not myopia risk factors in our study sample. In addition, we investigated whether chronotype affected myopia development and found that in our study sample, chronotype, assessed with MEQ, was not associated with myopia.

## Data Availability

All data produced in the present study are available upon reasonable request to the authors.

## List of abbreviations

CI: confidence interval
D: dioptre
MEQ: Morningness-Eveningness Questionnaire
OR: odds ratio
SD: standard deviation
SER: spherical equivalent refraction

## Declarations

### Ethics approval and consent to participate

Tallinn Medical Research Ethics Committee gave ethical approval for this work. Written informed consent was obtained from the participants and their parents or legal guardians.

### Consent for publication

Not applicable

